# Estimating the Number of Self-Harm Events Not Identified by Encounter Diagnoses in Health Records

**DOI:** 10.1101/2020.09.24.20200998

**Authors:** Gregory E Simon, Susan M Shortreed, Jennifer M Boggs, Gregory N Clarke, Rebecca C Rossom, Julie E Richards, Arne Beck

## Abstract

**Background:** Public health and research assessment of self-harm or suicide attempts must sometimes rely on health system records. Previous research regarding accuracy of encounter or billing diagnoses to identify self-harm or suicide attempt has yielded mixed results.

**Methods:** Using data from a large pragmatic trial of outreach to prevent suicide attempt, investigators identified ICD-10-CM diagnostic codes in categories where suicide attempt or self-harm might be missed or misclassified: injuries and poisonings coded as having “undetermined intent, injuries and poisonings coded as accidental, and injuries with no coding of intent. For injury and poisoning events with these selected codes, relevant text was extracted from full-text electronic health records and rated by a blinded panel of investigators as indicating or not indicating self-harm

**Results:** Diagnostic codes selected for review include 43 codes for injuries and poisonings with undetermined intent, 26 codes for accidental injuries and poisonings, and 46 codes for injuries without coding of intent. Among injury and poisoning events in this sample, full-text records were available for review for 60 coded as undetermined intent, 173 coded as accidents, and 288 with no coding of intent. After review of text, raters judged 46 (77%), 152 (88%), and 275 (95%) respectively as having adequate text documentation for confident rating of self-harm intent. Among those with adequate text for confident rating, the proportions of events classified as due to self-harm were 19% for injuries and poisonings coded as undetermined intent, 6% for injuries and poisonings coded as accidents, and 15% for injuries without coding of intent.

**Conclusions:** In this sample of people at high risk for self-harm or suicide attempt, we low rates of misclassified self-harm events among injuries and poisonings coded as having undetermined intent, accidental intent, or no coding of intent. In this sample, inclusion of all misclassified events identified through this search process would increase the total number of self-harm events identified by approximately 10%.

---

Public health and research assessment of self-harm or suicide attempts must sometimes rely on health system records. Population-level surveillance typically depends on insurance claims or billing records from emergency department and inpatient treatment settings. National suicide prevention efforts recommend use of records data to monitor progress as a core improvement strategy^1^. Evaluations of clinical strategies or care improvement efforts also rely on health records data as a primary or secondary source to evaluate clinical impact.

Previous research regarding accuracy of encounter or billing diagnoses to identify self-harm or suicide attempt has yielded mixed results^2^. In the ICD-9 diagnostic system in use until 2015, coding of intent (i.e. accident, assault, self-inflicted) required use of a separate cause-of-injury code or E-code. We have previously reported that actual use of those cause-of-injury codes varied widely across health systems and within health systems over time^3^. Among studies reviewing clinical notes for encounters with recorded diagnoses of self-inflicted injury or poisoning^4-6^, rates of documentation of self-harm ranged between 36% and 100% (i.e. false positive error rates from 0% to 64%). Assessments of sensitivity or false negative error rates have used variable methods and have reported widely varying estimates of the proportion of self-harm events not captured by ICD-9-CM codes^2^. These disparate findings regarding accuracy of coding likely reflect both true differences between settings in coding practices and differences in methods used to assess accuracy. We have previously reported that review of clinical notes for injuries and poisonings receiving ICD-9-CM diagnoses of “undetermined intent” often included clear documentation of self-harm^7^. Not surprisingly, varying findings have led to varying recommendations regarding use of ICD-9-CM diagnostic codes to identify self-harm^2, 7^.

Classification of injuries and poisonings changed significantly with the transition from the ICD-9-CM diagnostic system to ICD-10-CM. Coding of intent (i.e. accident, assault, or self-harm) was integrated into primary coding of all poisonings and some injuries, so that separate cause-of-injury codes are no longer required. The ICD-10-CM system also requires much more detailed specification of injury and poisoning categories. Using records data from several large integrated health systems, we have previously reported that this transition led to a marked decrease in rates of injuries and poisonings coded as having undetermined intent and a corresponding increase in rates of injuries and poisonings coded as self-harm^8^. We are aware of no research assessing accuracy of self-harm coding (either false positive or false negative errors) following the transition to ICD-10-CM.

We describe here a systematic assessment of potential false-negative errors in ICD10-CM diagnoses of self-harm extracted from health system records. Specifically, we use clinical notes to identify injury or poisoning self-harm events not coded as self-harm. We focus on three diagnostic categories in which misclassified self-harm events would most likely occur: injuries and poisonings coded as due to accidents, injuries and poisonings coded as having undetermined intent, and injuries classified as to site and type of injury with no coding of mechanism or intent. Findings regarding the accuracy of ICD-10-CM coding for self-harm should be useful for health systems evaluating the impact of suicide prevention efforts and researchers using records data to identify self-harm events.

## METHODS

This work was conducted to support a large pragmatic trial of outreach programs to prevent suicide attempt or other self-harm, conducted in four large health systems (HealthPartners and the Colorado, Northwest, and Washington regions of Kaiser Permanente). Trial methods are described in detail elsewhere^9, 10^ and are briefly summarized here. Between March 2015 and September 2018, adult outpatients seen in health system primary care or mental health clinics and reporting frequent “thoughts that you would be better off dead, or thoughts of hurting yourself in some way” on routinely administered depression questionnaires^11^ were automatically enrolled and randomly assigned to one of three intervention groups: one receiving continued usual care, one receiving usual care supplemented by an outreach and care management program, and one receiving usual care supplemented by a coach-supported online program teaching Dialectical Behavior Therapy (DBT)^12^ skills.

For each enrolled participant, health system electronic health record and insurance claims data identified all encounters in any setting (outpatient, emergency department, or inpatient) with any injury or poisoning diagnosis during 18 months after each participant’s date of randomization, including encounters at external facilities identified by insurance claims. Injury and poisoning codes occurring in this sample were sorted into three groups for potential record review. The first group, codes indicating undetermined intent, included all injury codes in the range of Y21 through Y33 as well as codes for poisoning (T36 through T65) and asphyxiation (T71) that included a specifier for undetermined intent.

The second group, c odes indicating accidental intent, included codes for poisoning (T36 through T65) and asphyxiation (T71) that included a specifier for accidental intent. The third group, Injury codes without coding of intent, included codes in the range from S00 through T32 that were not accompanied by an external cause code in the range from V00 through Y99.

All ICD-10-CM codes occurring in this sample in these three groups were then rated by a panel of five trial investigators (GS, RR, AB, GC, JB) with experience in use of health records data for suicide epidemiology and prevention research. Given our previous finding that codes for undetermined intent were often accompanied by documentation of self-harm, we presumed a relatively high rate of true self-harm events in that group. Consequently, investigators were asked to identify specific codes in the undetermined intent group that were unlikely to represent self-harm so that those could be excluded from chart review. Codes identified as unlikely mechanisms of self-harm by at least three investigators were excluded from further review, with all other codes in that group included. Conversely, we presumed a relatively low rate of true self-harm events among injuries or poisonings coded as accidents or injuries without coding of intent. Consequently, investigators were asked to identify specific codes in those groups indicating common or likely mechanisms of self-harm. Those identified as common mechanisms of self-harm by at least three investigators were selected for further review, with all other codes in those groups excluded.

The resulting code lists were then used to select specific injury or poisoning events for review of full-text clinical notes. Reviewable events were defined by occurrence of at least one reviewable diagnosis code (as described above) if there was no diagnosis code indicating definite self-harm occurring on the same day or any previous day (e.g. an injury or poisoning event with diagnostic codes for both self-harm and undetermined intent on the same day was not reviewed). This scheme was consistent with the study aim of measuring time to first self-harm event after randomization. A single injury or poisoning event could be selected based on diagnosis codes in two different groups (e.g. a code for poisoning of undetermined intent and a code for injury without coding of intent occurring on the same day). To preserve blinding regarding intervention group assignment, this review process was separated into two steps: extraction of relevant text by chart abstractors (not involved in intervention delivery) at each site followed by classification of that extracted and de-identified text by a separate panel of raters blinded to intervention group assignment and study site.

Chart abstractors were instructed to consider records of any encounters (outpatient, emergency department, inpatient, and telephone encounters) within 14 days before or after the date of the selected diagnosis. Abstractors identified and copied any text during that time period relevant to the intent of the injury or poisoning selected for review, including nursing notes, treating clinicians’ notes, and direct quotes from patients. Abstractors were advised to specifically identify text that would clarify presence or absence of self-harm intent, including both suicidal intent and intentional self-harm not necessarily accompanied by intent to die (i.e. non-suicidal self-injury). Abstraction typically began with any encounter on the diagnosis data, extending to encounters before and after that date until clear documentation of intent was identified or until all encounters during the interval were reviewed. Abstractors redacted any information that might reveal study intervention group assignment, any information that might allow re-identification of individual patients, and any information that might identify healthcare providers or facilities.

All relevant text for each event were then presented to a panel of six study investigators, with each event considered by three raters and each rater considering approximately half of all events. Raters were instructed to rate each injury/poisoning event as self-harm (a forced-choice rating of yes or no) and a separate three-level (high/medium/low) rating of confidence in that self-harm classification.

Descriptive analyses of ratings examined the distribution of confidence ratings and agreement among reviewers regarding self-harm classification. Those analyses informed selection of a threshold of confidence ratings. Subsequent descriptive analyses examined the distribution of rater’s self-harm classifications limited to events with adequate ratings of confidence.

Responsible institutional review boards at each participating health system reviewed and approved all trial procedures, including the chart review work described here.

## RESULTS

Among all injury and poisoning diagnoses recorded in the study sample during the follow-up period, records data included 50 ICD-10 CM diagnosis codes for injuries or poisonings with undetermined intent, 94 codes for injuries or poisonings coded as accidental, and 3702 codes for injuries not accompanied by coding of intent. Among undetermined intent codes, the panel of investigators classified 43 as plausible mechanisms of self-harm (i.e. included in review) and classified 7 as unlikely mechanisms for self-harm (i.e. excluded from review). Among accidental injury/poisoning codes, the panel of investigators identified 26 as common mechanisms of self-harm (i.e. included in review). Among injury codes with no coding of intent, the panel of investigators identified 46 as common mechanisms of self-harm (i.e. included in review). The most frequent included and excluded codes in each group are shown in Table 1, and a complete list is available from the corresponding author on request.

**Table 1.** Most common injury and poisoning diagnosis codes selected for inclusion and exclusion

|  | Included in record review | Excluded from record review |
| --- | --- | --- |
| **Undetermined Intent** | T50.904A – Poisoning by unspecified drug, undetermined intent<br>T42.4X4A – Poisoning by benzodiazepine, undetermined intent<br>T65.94XA – Toxic effect of unspecified substance, undetermined intent<br>T51.94XA – Toxic effect of unspecified alcohol, undetermined intent<br>T43.594A – Poisoning by antipsychotics, undetermined intent | T63.304A – Toxic effect of spider venom, undetermined intent<br>T63.444A – Toxic effect of bee venom, undetermined intent<br>T63.464A – Toxic effect of wasp venom, undetermined intent<br>T59.3X4A – Toxic effect of lacrimogenic gas, undetermined intent<br>T63.484A – Toxic effect of other arthropod venom, undetermined intent |
| **Accidental** | T42.4X1A – Poisoning by benzodiazepine, accidental<br>T43.591A – Poisoning by antipsychotic, accidental<br>T40.2X1A – Poisoning by other opioid, accidental<br>T40.601A – Poisoning by unspecified narcotic, accidental<br>T42.6X1A – Poisoning by antiepileptic or sedative/hypnotic, accidental | T50.901A – Poisoning by unspecified drug, accidental<br>T63.441A – Toxic effect of bee venom, accidental<br>T63.481A – Toxic effect of other arthropod venom, accidental<br>T56.891A - Toxic effect of other metals, accidental<br>T63.461A – Toxic effect of wasp venom, accidental |
| **No Coding of Intent** | S51.812A – Laceration without foreign body of left forearm<br>S51.811A - Laceration without foreign body of right forearm<br>S61.512A - Laceration without foreign body of left wrist<br>S61.511A – Laceration without foreign body of right wrist<br>S51.802A – Unspecified open wound of left forearm | S09.90XA – Unspecified injury of head<br>S39.012A – Strain of muscle, fascia, and tendon of lower back<br>S16.1XXA – Strain of muscle, fascia, and tendon of neck<br>T14.8XXA – Other injury of unspecified body region<br>S93.401A – Sprain of unspecified ligament of right ankle |

The selected list of undetermined intent codes identified 156 injury or poisoning events in this patient sample. Of those events, 70 had a definite self-harm code recorded on the same day, and 14 had a self-harm code recorded on a previous day, leaving 72 undetermined intent events to be reviewed. The selected list of accidental intent codes identified 308 injury or poisoning events in this patient sample. Of those events, 63 events had a definite self-harm code recorded on the same day, and 26 had a definite self-harm code recorded on a previous day, leaving 219 accidental events to be reviewed. The selected list of injury codes without coding of intent identified 499 injury events in this sample. Of those events, 121 had a definite self-harm code recorded on the same day, and 62 had a definite self-harm code recorded on a previous day, leaving 316 injury events without coding of intent to be reviewed.

Review of clinical notes for all encounters within 14 days before or after each of those events found no relevant text (i.e. no encounters with any mention of injury or poisoning) in 12 (17%) of undetermined intent events, 46 (21%) of accidental intent events, and 28 (9%) of injury events without coding of intent. Exclusion of those events left 60 events receiving a code for undetermined intent, 173 events receiving a code for accidental intent, and 288 injury events with no coding of intent. Because some events received codes in more than one category, the total number of unique events was 508.

Table 3 displays the distribution of confidence ratings and inter-rate agreement in each diagnosis group for events in with any relevant clinical text available for review. Raters expressed high confidence in ratings (summed confidence score of 6 or higher) for 77% of events coded as undetermined intent, 88% of events coded as accidents, and 95% of injuries without coding of intent. Agreement among reviewers was consistent with those confidence ratings; classification of self-harm was unanimous for the vast majority of events with high confidence ratings and a minority of events with low confidence ratings.

**Table 2.**
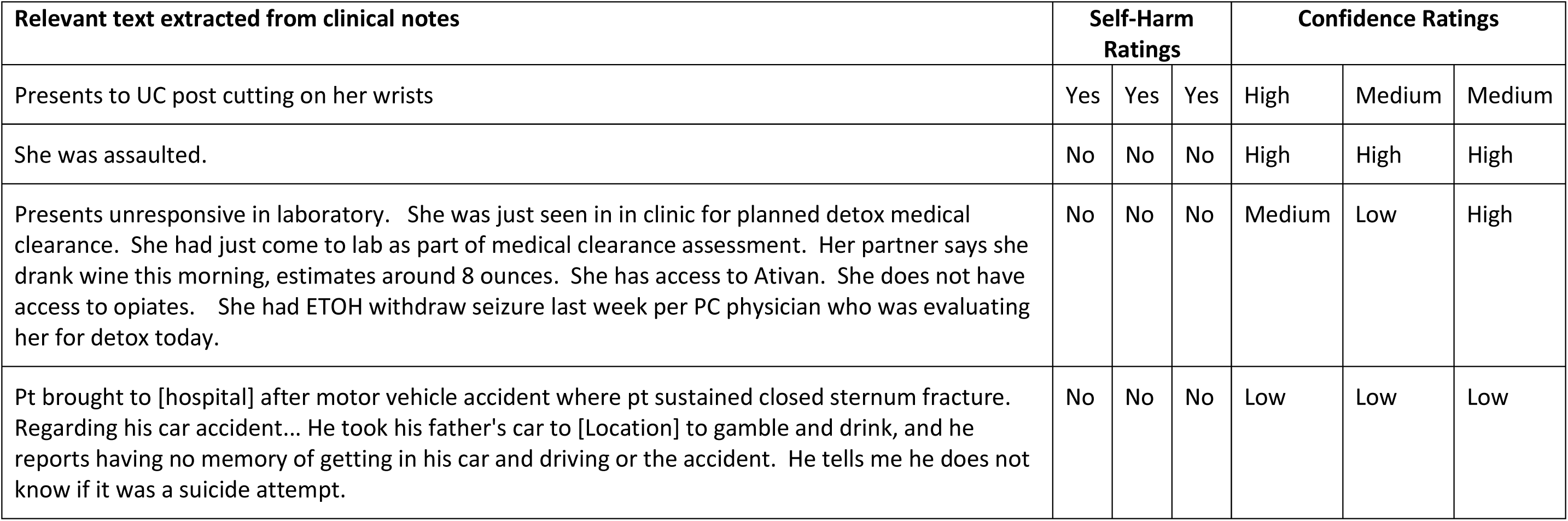
Examples of abstracted text and rater classifications

**Table 3.** Distribution of rater confidence and inter-rater agreement. Confidence scores calculated as sum of three individual rater scores where 1=low confidence, 2=medium confidence, and 3=high confidence.

|  | Coded As Undetermined Intent |  | Coded as Accidental Intent |  | Injury with No Coding of Intent |  |
| --- | --- | --- | --- | --- | --- | --- |
| Summed Confidence score | Number Total=60 | All three reviewers agreed N (%) | Number Total=173 | All three reviewers agreed N (%) | Number Total=288 | All three reviewers agreed N (%) |
| 3 | 5 | 2 (40%) | 7 | 1 (14%) | 3 | 1 (33%) |
| 4 | 3 | 1 (33%) | 6 | 1 (17%) | 5 | 4 (80%) |
| 5 | 6 | 2 (33%) | 8 | 5 (63%) | 5 | 3 (60%) |
| 6 | 13 | 10 (77%) | 23 | 18 (78%) | 12 | 9 (75%) |
| 7 | 6 | 6 (100%) | 32 | 28 (88%) | 22 | 18 (81%) |
| 8 | 11 | 11 (100%) | 29 | 28 (97%) | 36 | 34 (94%) |
| 9 | 16 | 16 (100%) | 68 | 67 (99%) | 205 | 204 (99%) |

Table 4 displays the distribution of self-harm ratings in each of the three diagnosis groups, limited to events with confidence scores of six or greater. In none of the groups, even with high confidence ratings, was the proportion of events with a majority of yes ratings higher than 19%. The total number of events with summed confidence score of six or greater and at least two reviewers classifying as self-harm was 58, including 9 events originally coded as undetermined intent, 8 originally coded as accidents, and 41 without coding of intent.

**Table 4.** Distribution of self-harm ratings, limited to events with text available and summed confidence score of 6 or higher.

|  | All 3 No N (%) | 2 No / 1 Yes N (%) | 1 No / 2 Yes N (%) | All 3 Yes N (%) |
| --- | --- | --- | --- | --- |
| Undetermined Intent | 36(78%) | 1 (2%) | 2 (4%) | 7 (15%) |
| Accidental Intent | 137 (90%) | 7 (5%) | 4 (3%) | 4 (3%) |
| No Intent Coding | 233 (85%) | 1 (0.4%) | 9 (3%) | 32 (12%) |

## DISCUSSION

Findings in this sample are generally reassuring regarding false-negative errors in the use of ICD-10-CM encounter diagnoses to identify health care encounters for self-harm. In the three groups of diagnoses examined, between 5 and 15% of injury and poisoning encounters had clinical text documenting intent of self-harm. In this sample, inclusion of all additional events identified by chart review would increase the total number of self-harm events identified by approximately 10% (58 events added to approximately 550 events coded as definite self-harm). If the rates observed among encounters with adequate text to review were extrapolated to the remaining encounters without adequate text, that proportion of additional events would increase to only 12%.

The low rates of self-harm we observed are notable given the selection of this sample. First, trial participants were selected for elevated risk of self-harm or suicidal behavior^9^. Our previous research suggests that outpatients reporting frequent thoughts of self-harm or suicide have a risk of subsequent self-harm more than ten times as high as the general population of health plan members^11, 13^. We would expect lower rates of self-harm in a population with a lower prior probability or expected prevalence^14^. Second, review was limited to diagnostic codes with higher likelihood of self-harm, eliminating injuries and poisonings unlikely to represent self-harm (among codes for undetermined intent) and selecting diagnostic codes more likely to represent self-harm (among codes for accidental intent and injuries with no coding of intent). We would expect even lower rates of self-harm events in these coding categories without this selection for codes more likely to represent self-harm.

Findings in this sample regarding injury and poisoning diagnoses coded as having undetermined intent differed markedly from our previous findings regarding ICD-9-CM coding of undetermined intent^7^. If limited to people with recent mental health treatment, approximately 80% of injury and poisoning events with ICD-9-CM diagnoses of undetermined intent had clear documentation of self-harm. The discrepancy between these new results and our previous results regarding undetermined intent diagnoses may be explained by the marked decrease in undetermined intent diagnoses we observed with the transition from ICD-9-CM to ICD-10-CM^8^. If the transition to the more specific ICD-10-CM coding scheme led to increased coding of self-harm, then the events remaining in the undetermined intent category might be less likely to represent self-harm or suicide attempt. Our previous findings and these findings indicate that monitoring or comparison of self-harm rates across the transition from ICD-9-CM to ICD-10-CM would be seriously flawed.

We are not aware of other data regarding prevalence of self-harm among injuries and poisonings coded as accidental or among injuries without coding of intent. Among injuries and poisonings coded as accidental, we found that only 5% were re-classified as self-harm after chart abstraction and blinded classification. Consequently, this group contributed only a small number of additional events. Among injuries with no coding of intent, we found that almost 15% were re-classified as self-harm. Given the relatively large number of injuries without coding of intent, this group contributed most of the additional events identified by chart review.

We should emphasize that this work does not consider many sources of false-negative error in use of encounter diagnoses to identify self-harm. First, people experiencing self-harm might not seek medical care, so no encounter would appear in health system records. Second, treating providers might not record an injury or poisoning diagnosis. Third, some self-harm events might receive non-specific diagnoses not included in our record review process.

Because we aimed to identify the first self-harm event among participants in this trial, we did not review injuries and poisonings occurring after a clear diagnosis of self-harm. We might observe different rates of misclassified self-harm events if we considered all potential events for any individual patient.

We should also caution that our findings may not generalize to other health systems. While the ICD-10-CM taxonomy now applies across the US, recording of encounter or billing diagnoses is certainly subject to local influences^3^. Diagnoses from outpatient and emergency department encounters, typically recorded by treating clinicians, are influenced by how possible codes are presented and prioritized within the facility or health system EHR environment. Diagnoses from inpatient encounters, sometimes recorded by treating clinicians and sometimes by designated coding consultants, are influenced by local policies, practices, and EHR environments.

Allowing for uncertainty regarding generalizability of these findings, we suggest the following implications. Researchers aiming to use health system data for population-based research, such as development of suicide risk prediction models, should not generally include diagnoses of undetermined intent in definitions of self-harm. Health systems aiming to measure or monitor population-level rates of self-harm should recognize that ICD-10-CM diagnoses of self-harm will modestly under-estimate true incidence rates.

**Supplementary Table A.** Number of times the same event was assigned diagnosis codes from multiple code categories on the same day.

|  | Definite self-harm code | Undetermined Intent code selected for review | Accidental code selected for review | Injury with no intent coding selected for review |
| --- | --- | --- | --- | --- |
| Definite self-harm code | N/A | 70 | 63 | 121 |
| Undetermined Intent code selected for review | 70 | N/A | 31 | 12 |
| Accidental code selected for review | 63 | 31 | N/A | 10 |
| Injury with no intent coding selected for review | 121 | 12 | 10 | N/A |

**Supplementary Table B.** Distribution of events receiving diagnosis codes in more than one review category, limited to events with relevant text available for review.

|  | Undetermined Intent code selected for review | Accidental code selected for review | Injury with no intent coding selected for review |
| --- | --- | --- | --- |
| Undetermined Intent code selected for review | N/A | 12 | 1 |
| Accidental code selected for review | 12 | N/A | 1 |
| Injury with no intent coding selected for review | 1 | 1 | N/A |

## Data Availability

We are not able to share clinical text from patient records.

## Notes

Supported by National Institute of Mental Health Cooperative Agreement UH3 MH007755

### Competing Interest Statement

The authors have declared no competing interest.

### Clinical Trial

NCT02326883

### Funding Statement

Supported by National Institute of Mental Health Cooperative Agreement UH3 MH007755.

### Author Declarations

All trial procedures, including the chart review work described here, were reviewed and approved by institutional review boards at each of the four participating health systems (HealthPartners IRB, Kaiser Permanente Colorado IRB, Kaiser Permanente Northwest IRB, and Kaiser Permanente Washington IRB). All four IRBs approved a modified or abbreviated consent procedure for participation in the pragmatic trial and a waiver of consent for the use of records data described in this report.

